# Burden of Liver Cancer in China from 1990 to 2019 and projections to 2044: Findings from the Global Burden of Disease Study

**DOI:** 10.1101/2023.08.07.23293756

**Authors:** Jianglong Han, Chao Chen, Tian Tang, Wenmin Liu, Ruyan Chen, Si Li, Haiyu Deng, Tingting Jian, Liang Zhao, Zhenming Fu

## Abstract

**Background:** China has the highest liver cancer burden in the world. Prediction and comparison of the future trends of liver cancer in China and some representative areas may guide further control action.

**Methods:** Using data from the Global Burden of Disease Study, we assessed incidence, mortality, and disability-adjusted life-years of liver cancer in Mainland China, with reference to representative East Asia areas (Taiwan China, Japan, and Korea) and Western areas (the United Kingdom and the United States). The burden of liver cancer was evaluated and predicted using NOREPRED model from 1990 to 2044.

**Results:** Overall, the liver cancer incidence (28.1 to 10.6 per 100,000) and mortality (27.5 to 9.7 per 100,000) decreased from 1990 to 2015 in Mainland China, which were consistent with the trends of Eastern Asia areas. However, the disease burden in Mainland China were then plateaued and started to increase during 2015-2044 (10.6 to 14.8 per 100,000 for incidence; 9.7 to 14.02 per 100,000 for mortality), including the hepatitis-related liver cancer incidence (increase from 8.6 to 11.7 per 100,000). While the changing patterns of alcohol- and nonalcoholic steatohepatitis (NASH)- related liver cancer incidence were found similar among Mainland China (0.93 to 1.51 per 100,000 for alcohol; 0.5 to 0.73 per 100,000 for NASH) and Western countries in our projection.

**Conclusion:** The liver cancer burden in Mainland China is unexpectedly predicted to increase again after decades of decline. Future efforts must be made to resolve both the remaining hepatitis-related cancer burden and the changing etiologies.

## Introduction

Liver cancer is the sixth most common malignancy and the third cause of cancer-related death worldwide, with approximately 905,677 new cases and 830,180 deaths in 2020(1). The disease burden of liver cancer has increased globally over the last few decades, with the highest disease burden observed in East Asia(2, 3), especially in Mainland China(1). In 2020, Mainland China had 410,038 newly diagnosed liver cancer cases and 391,105 related deaths, which comprised half the global burden(1). Fortunately, attributed to great public health efforts in controlling hepatitis virus infection, the incidence and mortality rates of liver cancer were reported to be declining in East Asia in recent studies(4, 5).

Hepatitis B virus (HBV) and hepatitis C virus (HCV) infection are the main risk factors for liver cancer development, accounting for almost half of the cases worldwide(6). Currently, HBV infection is still the leading risk factor for liver cancer in China, while HCV infection is the major risk factor for liver cancer in South Korea and Japan and other developed areas(7, 8). However, liver cancer is a multifactorial disease(9). Other modifiable factors such as obesity and unhealthy lifestyles, including alcohol consumption, intake of fatty food, and lack of exercise, also play important roles in its development(10). Recently, it is reported that the high prevalence of alcoholic liver disease (ALD) and nonalcoholic fatty liver disease (NAFLD) or nonalcoholic steatohepatitis (NASH) in Western populations result in the upward trend in liver cancer (11, 12). The heterogeneous prevalence of risk factors in different regions may also lead to different disease burdens. In the past few decades, great efforts have been undertaken in East Asia, including Mainland China(13), to prevent hepatitis virus infection through national vaccination programs, blood supply management, and management of virus carriers. Therefore, the prevalence of hepatitis infection and related liver cancer burden was found to decline in this area(14–16).

Recently, there has been significant increase in the consumption of alcohol, refined grains, and highly processed foods, while physical activity decreased with increasing sedentary behaviors in Mainland China(17).As a result, the rapid emergence of alcohol- and NASH-related liver cancer may offset or even surpass the likely decline in the burden of hepatitis-related liver cancer in Mainland China. Therefore, we used the data derived from the Global Burden of Disease (GBD) Study 2019 to examine the current trends of liver cancer burden in China by specific etiologies and projected the trends to 2044.

## Materials and methods

### Data source

The GBD study provides a standardized approach for estimating incidence, mortality, and disability-adjusted life-years (DALYs), as well as DALYs by cause, age, sex, year, and location, aiming to use all accessible information on disease occurrence, natural history, and severity that passes a set of inclusion criteria(18). The detailed methods of the GBD study (1990–2019) have been reported in previous studies(18, 19). We collected annual liver cancer case data between 1990 and 2019 by sex, region (Mainland China, East Asia areas: Taiwan China, Japan, and Republic of Korea; Western areas: US and UK), etiology (alcohol use, HBV, HCV, and NASH), and age group (from under 5 to 85+ years in 5-year intervals) from the Institute for Health Metrics and Evaluation (http://ghdx.healthdata.org/gbd-results-tool). The data on incidence, mortality, and DALYS were chosen as measures in the inquiry tools. The age-standardized incidence (ASIR), mortality (ASMR), and DALY (ASDR) rates were adjusted by the World Health Organization standard population distribution (2000-2025) per 100,000 person-years. Patient consent was not required because the study is a retrospective database research in nature, and there was no direct patient contact. Institutional Review Board approval was not required according to our institution policy.

### Statistical analysis

We described the liver cancer incidence, mortality, and DALYs by 5-year age groups, locations, and etiologies. Temporal trends of these variables were plotted by location and etiology using the age-standardized rates. The temporal trends were divided into two periods: 1990-2019 was regarded as the observed trend plotted as solid lines, and 2020-2044 was regarded as the predicted trend plotted as dashed lines.

We used a log-linear age-period-cohort model to level off exponential growth and limit linear trend forecasts to observed trends (1990–2019). The NOREPRED package with Power5 and Poisson APC models in R software were developed and implemented in projecting trends into the future(20). We further utilized the APC method to make projections of liver cancer burden up to 2044. Projections were carried out in 5-year intervals through 2020-2044. Changing trends in observed periods (1990–2019) were extrapolated using the power function of the NOREPRED package to level off the growth, with a projection of the linear trends for the last two observed 5-year periods. The projected trend was attenuated by 25% and 50% for the second (2025-2029) and third (2030-2034) predicted periods, respectively, and then 75% for the fourth and fifth projected periods (2035-2044)(21). The predicted population by age group and region from 2020 to 2044 was obtained from the United Nations World Population Prospects 2019 Revisions in a 5-year period(21). R 4.1 was used for analysis, including the functions available in R studio and the NORDPRED package. GraphPad Prism 8 was used to visualize the results.

## Results

### Incidence, mortality, and DALYs of liver cancer by region, 1990-2019

Overall, the highest incidence, mortality, and DALY rates of liver cancer were observed in East Asia from 1990 to 2019 (**Figure 1A, 2A**, and **3A**). The highest ASIR was observed in Mainland China at 28.1 per 100,000 in 1990, followed by Japan at 16.16 per 100,000, Taiwan China, at 12.83 per 100,000, and South Korea at 9.14 per 100,000. Between 1990 and 2019, continuously decreased incidences were observed in Mainland China, Japan, and Taiwan China, whereas the incidence in South Korea increased 184% from 1990 to 2004 and then decreased from 23.63 to 11.44 per 100,000 through 2019. The UK (2.01 to 4.82 per 100,000) and US (2.36 to 4.94 per 100,000) showed slight but continuous increases in incidence over the period.

**Figure 1.**
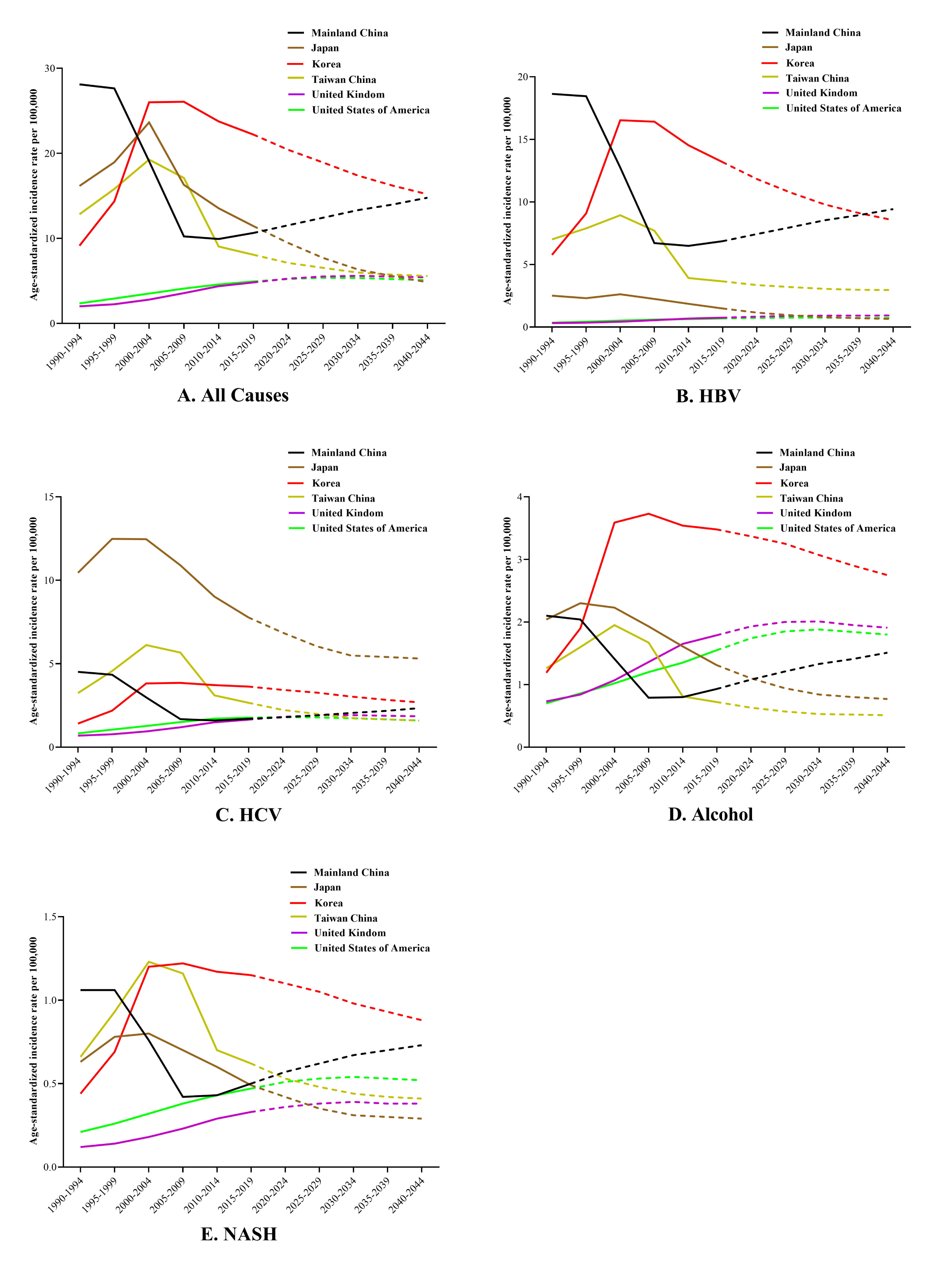
Trends in liver cancer incidence through 1990 to 2045: observed (solid lines) and predicted rates (dashed lines). Age-standardized to world population per 100,000.

Similarly, the highest ASMR and ASDR were observed in Mainland China, followed by Japan, Taiwan China, and South Korea in 1990, but significant decreasing trends were also observed in these areas. The figures in Mainland China decreased for more than two decades from 1990 to 2015 but started to increase again in 2015. However, unlike the trends of ASIR and ASDR, the temporal trends of South Korea decreased in the ASMR through 2019. Slight but steady increases in ASMR and ASDR were observed in the UK and US.

### Incidence, mortality, and DALYs of liver cancer by etiology, 1990-2019

Among all four underlying etiologies, the highest disease burden was observed in HBV-related liver cancer, followed by HCV, alcohol consumption, and NASH in China and selected areas (**Table 1**). Overall, the highest ASIRs were observed in Mainland China among HBV (18.63 per 100,000), alcohol (2.1 per 100,000), and NASH-related (1.06 per 100,000) liver cancer in 1990, whereas the highest HCV-related ASIR of liver cancer (10.45 per 100,000) was seen in Japan. Remarkable decreases were found in the ASIRs of alcohol (-62%), HBV (-64%), HCV (-63%), and NASH (-60%) related to liver cancer in Mainland China from 1990 to 2015, and ASIRs have plateaued or slightly increased since 2015 (**Table 2**). In contrast, the ASIR of South Korea showed a rapid increase from 1990 to 2004 among all etiology-related liver cancers and then slowly decreased through 2019. Similar increasing trends were observed in the US and UK for liver cancers due to alcohol and NASH, whereas the ASIRs of hepatitis virus-related liver cancer plateaued in the UK and US throughout the study periods. Similar trends for ASMR and ASDR were also found in all regions regardless of etiology, with the highest disease burden of hepatitis virus-related liver cancers in Eastern countries and consistent upward trends in alcohol and NASH-related disease in Western countries.

**Table 1.** Disease burden of liver cancer from GBD by etiologies, 1990-2044.

| Period | Mainland China |  |  | Taiwan China |  |  | Japan |  |  | Republic of Korea |  |  | United Kingdom |  |  | United States of America |  |  |
| --- | --- | --- | --- | --- | --- | --- | --- | --- | --- | --- | --- | --- | --- | --- | --- | --- | --- | --- |
|  | ASIR | ASMR | ASDR | ASIR | ASMR | ASDR | ASIR | ASMR | ASDR | ASIR | ASMR | ASDR | ASIR | ASMR | ASDR | ASIR | ASMR | ASDR |
| All causes |  |  |  |  |  |  |  |  |  |  |  |  |  |  |  |  |  |  |
| 1990-1994 | 28.1 | 27.49 | 845.3 | 12.83 | 12.65 | 373.88 | 16.16 | 12.22 | 312.74 | 9.14 | 9.24 | 263.76 | 2.01 | 1.81 | 44.38 | 2.36 | 2.06 | 54.23 |
| 1995-1999 | 27.63 | 27.63 | 839.76 | 15.81 | 12.21 | 347.64 | 18.94 | 15.25 | 370.6 | 14.34 | 12.28 | 349.67 | 2.26 | 1.98 | 48.34 | 2.93 | 2.47 | 65.04 |
| 2000-2004 | 19.07 | 17.17 | 507.85 | 19.26 | 12.72 | 343.03 | 23.63 | 14.07 | 324.66 | 25.99 | 23.75 | 669.45 | 2.8 | 2.35 | 56.44 | 3.51 | 2.89 | 76.32 |
| 2005-2009 | 10.23 | 9.89 | 284.41 | 17.1 | 13.25 | 345.99 | 16.29 | 11.71 | 258.2 | 26.05 | 20.31 | 552.93 | 3.56 | 2.91 | 69.25 | 4.1 | 3.36 | 88.85 |
| 2010-2014 | 9.93 | 8.87 | 252.97 | 9.04 | 7.23 | 181.28 | 13.53 | 9.35 | 198.1 | 23.74 | 17.49 | 456.51 | 4.37 | 3.48 | 80.63 | 4.58 | 3.69 | 96.32 |
| 2015-2019 | 10.64 | 9.65 | 276.52 | 8.07 | 6.72 | 168.4 | 11.44 | 7.88 | 162.55 | 22.2 | 15.71 | 400.04 | 4.82 | 3.74 | 86.45 | 4.94 | 4.01 | 103.95 |
| 2020-2024 | 11.54 | 10.6 | 308.92 | 7.11 | 6.15 | 155.24 | 9.45 | 6.66 | 136.28 | 20.4 | 13.82 | 345.28 | 5.27 | 3.98 | 91.81 | 5.23 | 4.27 | 109.21 |
| 2025-2029 | 12.43 | 11.53 | 338.64 | 6.53 | 5.8 | 146.62 | 7.7 | 5.66 | 117.6 | 18.92 | 12.44 | 305.04 | 5.52 | 4.08 | 94.67 | 5.36 | 4.4 | 111.16 |
| 2030-2034 | 13.32 | 12.47 | 365.61 | 5.98 | 5.37 | 138.49 | 6.34 | 5.01 | 106.45 | 17.38 | 11.14 | 273.04 | 5.6 | 4.07 | 95.23 | 5.34 | 4.39 | 109.91 |
| 2035-2039 | 13.98 | 13.16 | 384.04 | 5.73 | 5.15 | 133.94 | 5.56 | 4.87 | 103.92 | 16.2 | 10.29 | 252.61 | 5.5 | 3.94 | 93.47 | 5.21 | 4.27 | 106.51 |
| 2040-2044 | 14.78 | 14.02 | 404.56 | 5.58 | 5.01 | 131.87 | 4.84 | 4.69 | 102.15 | 15.18 | 9.53 | 235.3 | 5.42 | 3.83 | 92.15 | 5.07 | 4.14 | 103.55 |
| Alcohol |  |  |  |  |  |  |  |  |  |  |  |  |  |  |  |  |  |  |
| 1990-1994 | 2.1 | 2.08 | 58.98 | 1.26 | 1.28 | 34.96 | 2.04 | 1.54 | 40.89 | 1.19 | 1.23 | 32.14 | 0.73 | 0.65 | 16.02 | 0.7 | 0.61 | 15.81 |
| 1995-1999 | 2.04 | 2.06 | 57.2 | 1.6 | 1.24 | 33.52 | 2.3 | 1.84 | 46.8 | 1.9 | 1.65 | 43.46 | 0.84 | 0.72 | 17.82 | 0.86 | 0.72 | 19.02 |
| 2000-2004 | 1.41 | 1.3 | 34.95 | 1.95 | 1.3 | 33.78 | 2.23 | 1.65 | 40.34 | 3.59 | 3.33 | 86.83 | 1.07 | 0.88 | 21.36 | 1.02 | 0.82 | 22.21 |
| 2005-2009 | 0.79 | 0.78 | 20.31 | 1.67 | 1.3 | 33.03 | 1.93 | 1.36 | 31.94 | 3.73 | 2.99 | 75.18 | 1.36 | 1.09 | 26.2 | 1.2 | 0.96 | 26 |
| 2010-2014 | 0.8 | 0.73 | 18.87 | 0.81 | 0.66 | 16.18 | 1.61 | 1.07 | 24.58 | 3.54 | 2.68 | 64.93 | 1.65 | 1.27 | 29.97 | 1.35 | 1.06 | 28.49 |
| 2015-2019 | 0.93 | 0.62 | 22.25 | 0.72 | 0.6 | 14.78 | 1.31 | 0.87 | 19.73 | 3.48 | 2.52 | 60.07 | 1.79 | 1.35 | 31.67 | 1.55 | 1.23 | 32.46 |
| 2020-2024 | 1.08 | 0.55 | 26.34 | 0.63 | 0.55 | 13.45 | 1.1 | 0.72 | 16.16 | 3.37 | 2.33 | 54.85 | 1.93 | 1.42 | 33.14 | 1.74 | 1.39 | 35.89 |
| 2025-2029 | 1.21 | 0.5 | 30.18 | 0.57 | 0.52 | 12.71 | 0.94 | 0.61 | 13.77 | 3.25 | 2.18 | 50.58 | 2 | 1.44 | 33.71 | 1.85 | 1.48 | 37.73 |
| 2030-2034 | 1.33 | 0.5 | 33.61 | 0.53 | 0.5 | 12.24 | 0.84 | 0.55 | 12.4 | 3.07 | 1.99 | 46.54 | 2.01 | 1.42 | 33.52 | 1.88 | 1.5 | 37.92 |
| 2035-2039 | 1.41 | 0.51 | 35.88 | 0.52 | 0.49 | 12.15 | 0.8 | 0.53 | 12.05 | 2.9 | 1.86 | 43.54 | 1.95 | 1.37 | 32.62 | 1.84 | 1.46 | 36.87 |
| 2040-2044 | 1.51 | 0.53 | 38.54 | 0.51 | 0.48 | 12.36 | 0.77 | 0.53 | 11.92 | 2.75 | 1.74 | 41.04 | 1.91 | 1.33 | 31.87 | 1.8 | 1.43 | 36.05 |
| HBV |  |  |  |  |  |  |  |  |  |  |  |  |  |  |  |  |  |  |
| 1990-1994 | 18.63 | 17.81 | 588.81 | 7.01 | 6.72 | 219.66 | 2.51 | 1.82 | 56.18 | 5.77 | 5.68 | 177.81 | 0.32 | 0.26 | 7.86 | 0.35 | 0.29 | 8.92 |
| 1995-1999 | 18.45 | 18.08 | 588.06 | 7.89 | 5.93 | 190.56 | 2.31 | 2.13 | 62.64 | 9.08 | 7.56 | 235.63 | 0.35 | 0.28 | 8.52 | 0.43 | 0.34 | 10.56 |
| 2000-2004 | 12.75 | 11.19 | 355.65 | 8.94 | 5.61 | 174.67 | 2.62 | 1.85 | 52.56 | 16.52 | 14.72 | 451.23 | 0.43 | 0.33 | 9.82 | 0.52 | 0.4 | 12.31 |
| 2005-2009 | 6.71 | 6.32 | 197 | 7.7 | 5.68 | 172.9 | 2.25 | 1.49 | 41.05 | 16.41 | 12.36 | 367.12 | 0.55 | 0.41 | 12.12 | 0.6 | 0.45 | 13.86 |
| 2010-2014 | 6.49 | 5.65 | 174.89 | 3.92 | 2.97 | 87.75 | 1.86 | 1.17 | 31.22 | 14.53 | 10.35 | 297.62 | 0.68 | 0.48 | 14.02 | 0.64 | 0.48 | 14.36 |
| 2015-2019 | 6.86 | 6.03 | 188.89 | 3.65 | 2.89 | 85.05 | 1.49 | 0.94 | 24.67 | 13.18 | 9.01 | 254.84 | 0.75 | 0.52 | 15.23 | 0.69 | 0.52 | 15.54 |
| 2020-2024 | 7.43 | 6.59 | 210.38 | 3.36 | 2.79 | 82.25 | 1.16 | 0.75 | 19.72 | 11.83 | 7.71 | 215.08 | 0.83 | 0.55 | 16.38 | 0.73 | 0.55 | 16.31 |
| 2025-2029 | 7.98 | 7.13 | 230.09 | 3.2 | 2.75 | 80.43 | 0.94 | 0.62 | 16.46 | 10.74 | 6.77 | 186.44 | 0.89 | 0.57 | 17.18 | 0.75 | 0.56 | 16.71 |
| 2030-2034 | 8.54 | 7.69 | 247.7 | 3.05 | 2.67 | 78.43 | 0.79 | 0.53 | 14.52 | 9.79 | 5.99 | 165.46 | 0.92 | 0.58 | 17.6 | 0.75 | 0.56 | 16.68 |
| 2035-2039 | 8.95 | 8.1 | 259.64 | 2.98 | 2.63 | 77.12 | 0.71 | 0.51 | 13.8 | 9.11 | 5.5 | 152.65 | 0.92 | 0.56 | 17.61 | 0.73 | 0.54 | 16.34 |
| 2040-2044 | 9.43 | 8.59 | 272.49 | 2.96 | 2.63 | 77.2 | 0.65 | 0.48 | 13.23 | 8.54 | 5.08 | 142.13 | 0.93 | 0.55 | 17.68 | 0.72 | 0.53 | 16.1 |
| HCV |  |  |  |  |  |  |  |  |  |  |  |  |  |  |  |  |  |  |
| 1990-1994 | 4.51 | 4.78 | 109.09 | 3.24 | 3.39 | 81.09 | 10.45 | 8 | 192.61 | 1.41 | 1.55 | 33.18 | 0.69 | 0.67 | 14.06 | 0.83 | 0.76 | 17.9 |
| 1995-1999 | 4.34 | 4.68 | 105.88 | 4.56 | 3.7 | 85.05 | 12.48 | 10.19 | 233.55 | 2.19 | 2.03 | 43.51 | 0.77 | 0.73 | 15.08 | 1.05 | 0.94 | 22.06 |
| 2000-2004 | 2.98 | 2.93 | 64.44 | 6.12 | 4.33 | 94.17 | 12.46 | 9.54 | 207.07 | 3.82 | 3.76 | 82.19 | 0.94 | 0.85 | 17.34 | 1.27 | 1.1 | 26.23 |
| 2005-2009 | 1.68 | 1.77 | 37.54 | 5.67 | 4.69 | 98.55 | 10.9 | 8.02 | 165.34 | 3.85 | 3.3 | 69.61 | 1.19 | 1.06 | 21.35 | 1.5 | 1.3 | 31.06 |
| 2010-2014 | 1.6 | 1.56 | 32.49 | 3.1 | 2.67 | 53.11 | 9.02 | 6.41 | 126.34 | 3.72 | 2.98 | 59.56 | 1.49 | 1.3 | 25.54 | 1.71 | 1.44 | 34.56 |
| 2015-2019 | 1.7 | 1.71 | 35.54 | 2.66 | 2.37 | 46.77 | 7.77 | 5.49 | 105.17 | 3.63 | 2.81 | 53.96 | 1.65 | 1.4 | 27.68 | 1.77 | 1.51 | 35.81 |
| 2020-2024 | 1.79 | 1.84 | 38.94 | 2.23 | 2.06 | 40.51 | 6.86 | 4.72 | 89.73 | 3.43 | 2.53 | 47.76 | 1.82 | 1.5 | 29.56 | 1.8 | 1.55 | 36.19 |
| 2025-2029 | 1.91 | 1.99 | 42.56 | 1.99 | 1.87 | 36.64 | 6.04 | 4.06 | 78.76 | 3.27 | 2.36 | 43.4 | 1.9 | 1.54 | 30.51 | 1.78 | 1.55 | 35.7 |
| 2030-2034 | 2.05 | 2.17 | 46.46 | 1.75 | 1.66 | 33.38 | 5.49 | 3.63 | 72.75 | 3.03 | 2.14 | 39.36 | 1.92 | 1.54 | 30.61 | 1.73 | 1.51 | 34.55 |
| 2035-2039 | 2.17 | 2.32 | 49.73 | 1.67 | 1.58 | 32.16 | 5.41 | 3.56 | 72.71 | 2.84 | 2.02 | 36.88 | 1.88 | 1.49 | 29.81 | 1.67 | 1.46 | 33.08 |
| 2040-2044 | 2.34 | 2.53 | 53.74 | 1.59 | 1.51 | 31.8 | 5.31 | 3.45 | 73.35 | 2.68 | 1.91 | 34.78 | 1.85 | 1.46 | 29.08 | 1.6 | 1.4 | 31.72 |
| **NASH** |  |  |  |  |  |  |  |  |  |  |  |  |  |  |  |  |  |  |
| 1990-1994 | 1.06 | 1.07 | 29.23 | 0.66 | 0.66 | 17.93 | 0.63 | 0.48 | 11.54 | 0.44 | 0.46 | 10.71 | 0.12 | 0.11 | 2.53 | 0.21 | 0.19 | 4.41 |
| 1995-1999 | 1.06 | 1.09 | 29.49 | 0.93 | 0.72 | 18.69 | 0.78 | 0.63 | 14.27 | 0.69 | 0.62 | 14.44 | 0.14 | 0.13 | 2.88 | 0.26 | 0.24 | 5.35 |
| 2000-2004 | 0.76 | 0.71 | 18.39 | 1.23 | 0.82 | 20.27 | 0.8 | 0.61 | 13.11 | 1.2 | 1.16 | 27.16 | 0.18 | 0.16 | 3.44 | 0.32 | 0.28 | 6.49 |
| 2005-2009 | 0.42 | 0.42 | 10.51 | 1.16 | 0.92 | 21.89 | 0.7 | 0.51 | 10.47 | 1.22 | 1.02 | 22.81 | 0.23 | 0.2 | 4.33 | 0.38 | 0.34 | 7.84 |
| 2010-2014 | 0.43 | 0.41 | 9.86 | 0.7 | 0.56 | 12.76 | 0.6 | 0.42 | 8.33 | 1.17 | 0.91 | 19.38 | 0.29 | 0.25 | 5.19 | 0.43 | 0.37 | 8.58 |
| 2015-2019 | 0.5 | 0.48 | 11.54 | 0.62 | 0.52 | 11.66 | 0.49 | 0.35 | 6.66 | 1.15 | 0.86 | 17.85 | 0.33 | 0.27 | 5.67 | 0.47 | 0.41 | 9.4 |
| 2020-2024 | 0.57 | 0.55 | 13.45 | 0.53 | 0.47 | 10.48 | 0.42 | 0.29 | 5.46 | 1.1 | 0.79 | 16.12 | 0.36 | 0.29 | 6.11 | 0.51 | 0.44 | 10.1 |
| 2025-2029 | 0.62 | 0.62 | 15.07 | 0.48 | 0.43 | 9.71 | 0.35 | 0.24 | 4.64 | 1.05 | 0.74 | 14.82 | 0.38 | 0.29 | 6.37 | 0.53 | 0.47 | 10.5 |
| 2030-2034 | 0.67 | 0.67 | 16.39 | 0.44 | 0.39 | 9 | 0.31 | 0.21 | 4.16 | 0.98 | 0.68 | 13.59 | 0.39 | 0.3 | 6.46 | 0.54 | 0.47 | 10.54 |
| 2035-2039 | 0.7 | 0.7 | 17.12 | 0.42 | 0.36 | 8.64 | 0.3 | 0.21 | 4.06 | 0.93 | 0.64 | 12.76 | 0.38 | 0.29 | 6.36 | 0.53 | 0.47 | 10.29 |
| 2040-2044 | 0.73 | 0.74 | 18.04 | 0.41 | 0.34 | 8.45 | 0.29 | 0.2 | 4 | 0.88 | 0.61 | 12.01 | 0.38 | 0.28 | 6.29 | 0.52 | 0.46 | 10.03 |

Abbreviation: GBD: Global Burden of Disease; HBV: hepatitis B virus; HCV: hepatitis C virus; NASH: non-alcoholic steatohepatitis; DALYs: Disability Adjusted Life Years

**Table 2.**
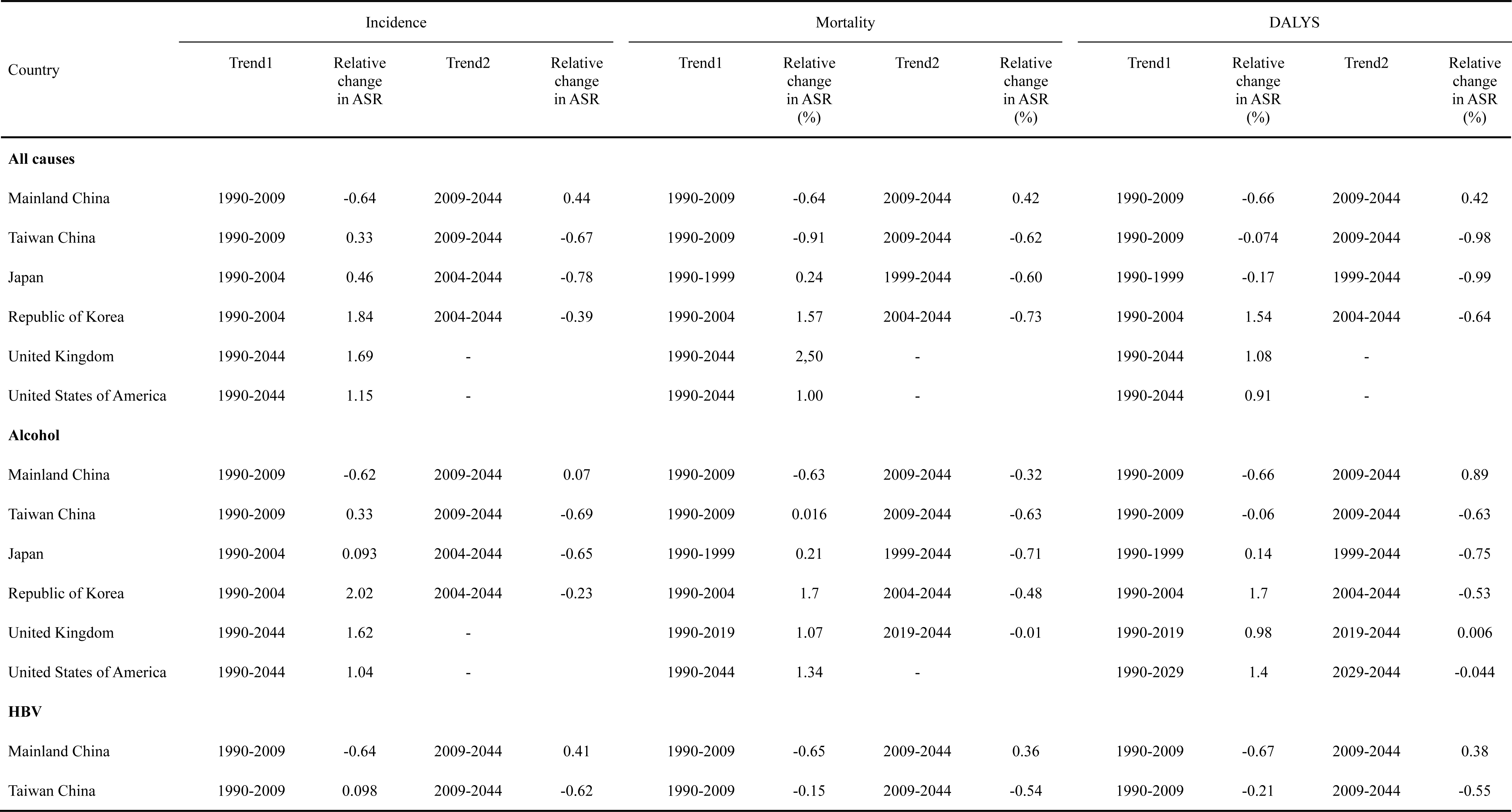
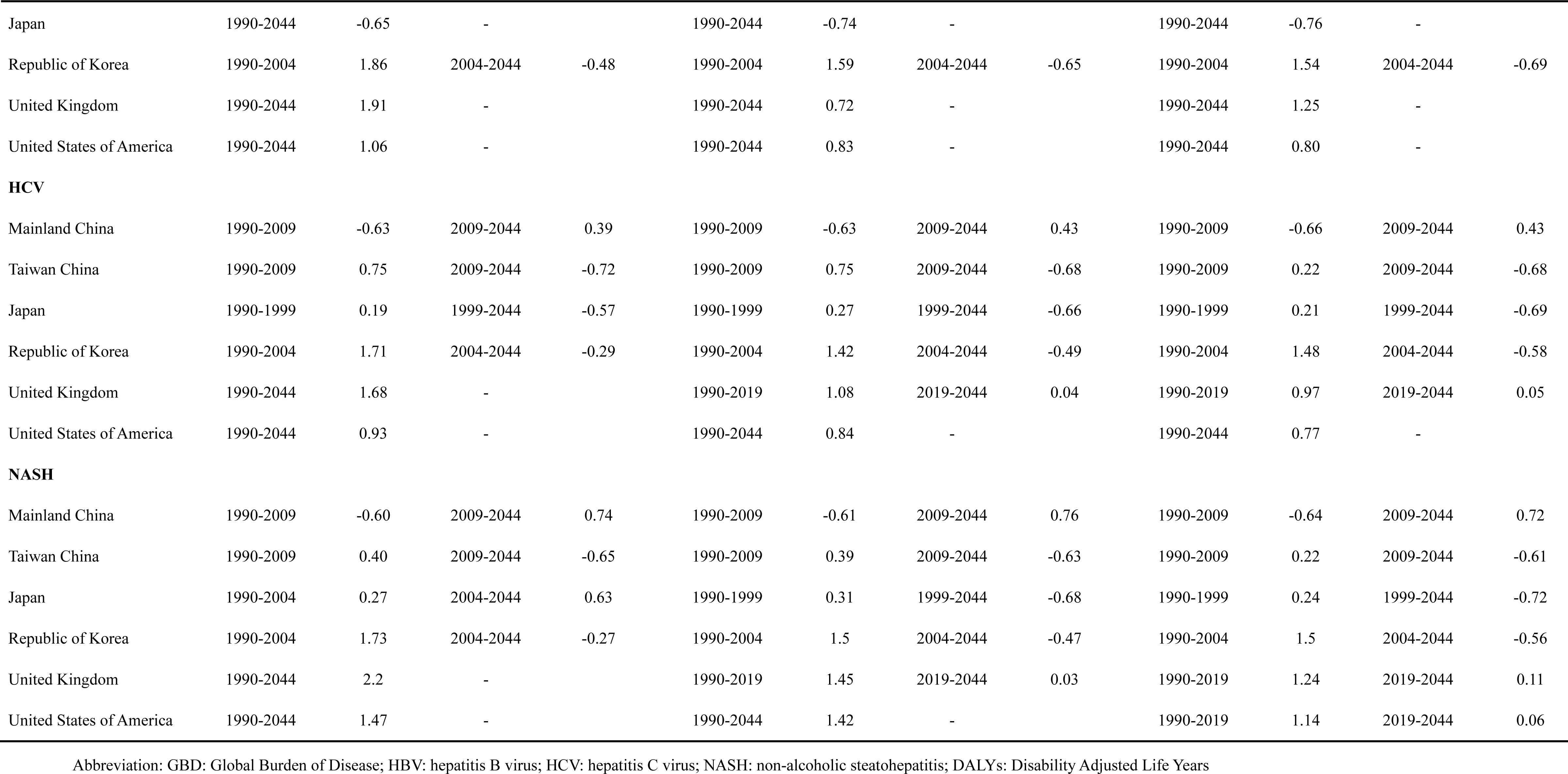
Trends in disease burden of liver cancer from GBD by etiologies, 1990-2044.

### Prediction of incidence, mortality, and DALYs of liver cancer, 2020-2044

Based on GBD data on liver cancer from 1990 to 2019, we further predicted the ASIR, ASMR, and ASDR in the next 25 years (**Figures 1, 2, 3**). By region, Mainland China, the US, and the UK showed continuous increases in incidence, mortality, and DALYs. The increases in Mainland China were 44% for incidence and 42% for mortality and DALYs, while the increases in incidence, mortality, and DALYs were 12.4%, 2.4%, and 6.6% for the UK and 2.6%, 3.2%, and 7.5% for the US, respectively (**Table 2**). In the rest of the regions, the incidence of liver cancer is predicted to decrease by 48.7% for Japan, 21.5% for Taiwan China, and 25.6% for Korea from 2020 to 2044, while mortality and DALYs in these areas will decline in a similar manner.

Universal increasing trends were observed for Mainland China for all etiological factors in incidence, mortality, and DALYs from 2020 to 2044. For HBV-related liver cancers, which are the predominant hepatitis virus-related liver cancers in Mainland China, trends were predicted to continuously increase in incidence (from 6.86 to 9.43 per 100,000), mortality (6.03 to 8.59 per 100,000), and DALYs (188.89 to 272.49 per 100,000) from 2020 to 2044. In Western areas, the incidence of liver cancer due to alcohol will increase by 6.7% for the UK and 16.1% for the US from 2020 to 2044, whereas NASH-related liver cancer increases by 15.1% for the UK and 10.6% for the US. However, mortality and DALYs will plateau in the UK and US over the predicted periods (**Figure 2D, E and Figure 3D, E**).

**Figure 2.**
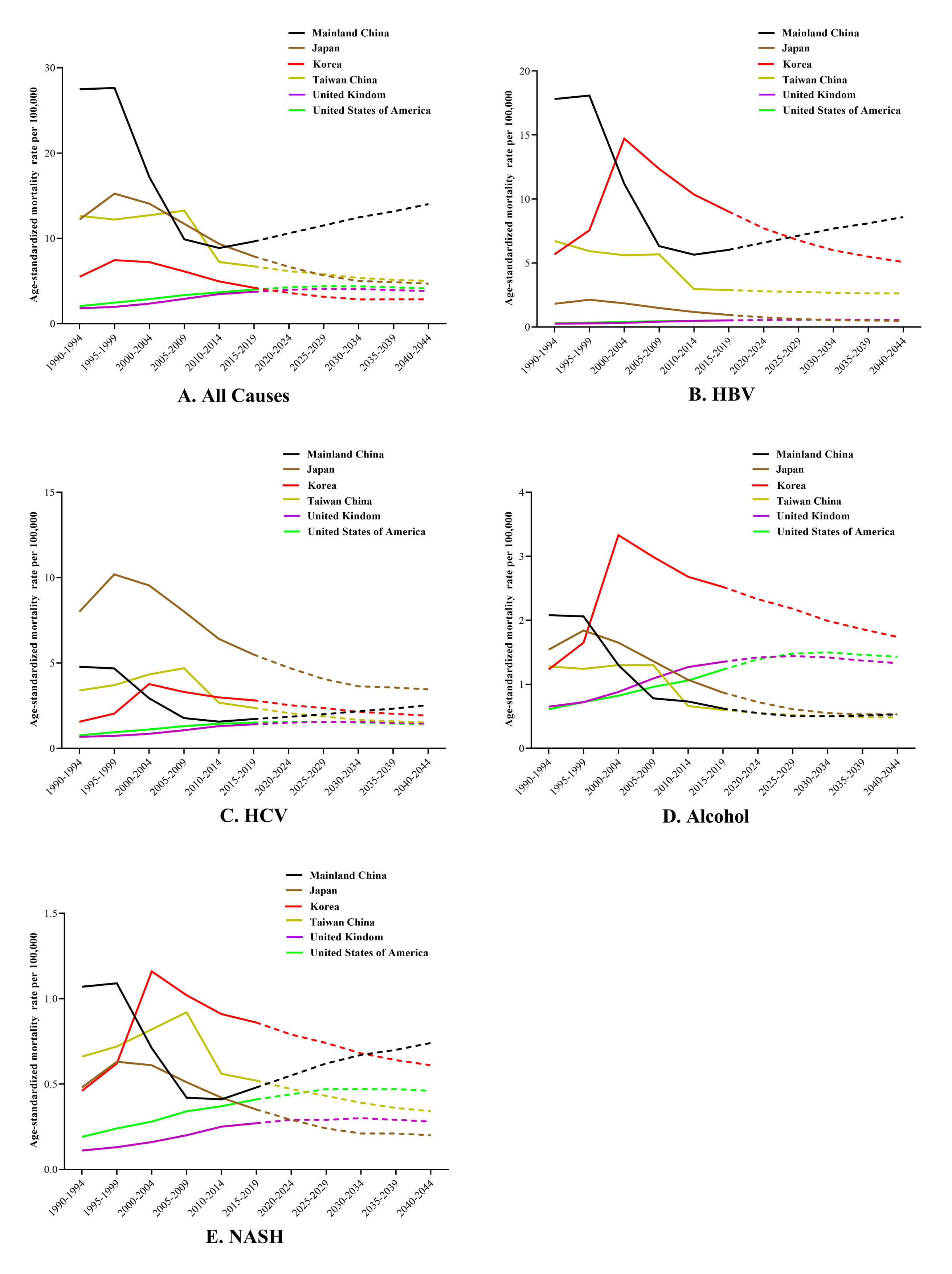
Trends in liver cancer mortality through 1990 to 2045: observed (solid lines) and predicted rates (dashed lines). Age-standardized to world population per 100,000.

**Figure 3.**
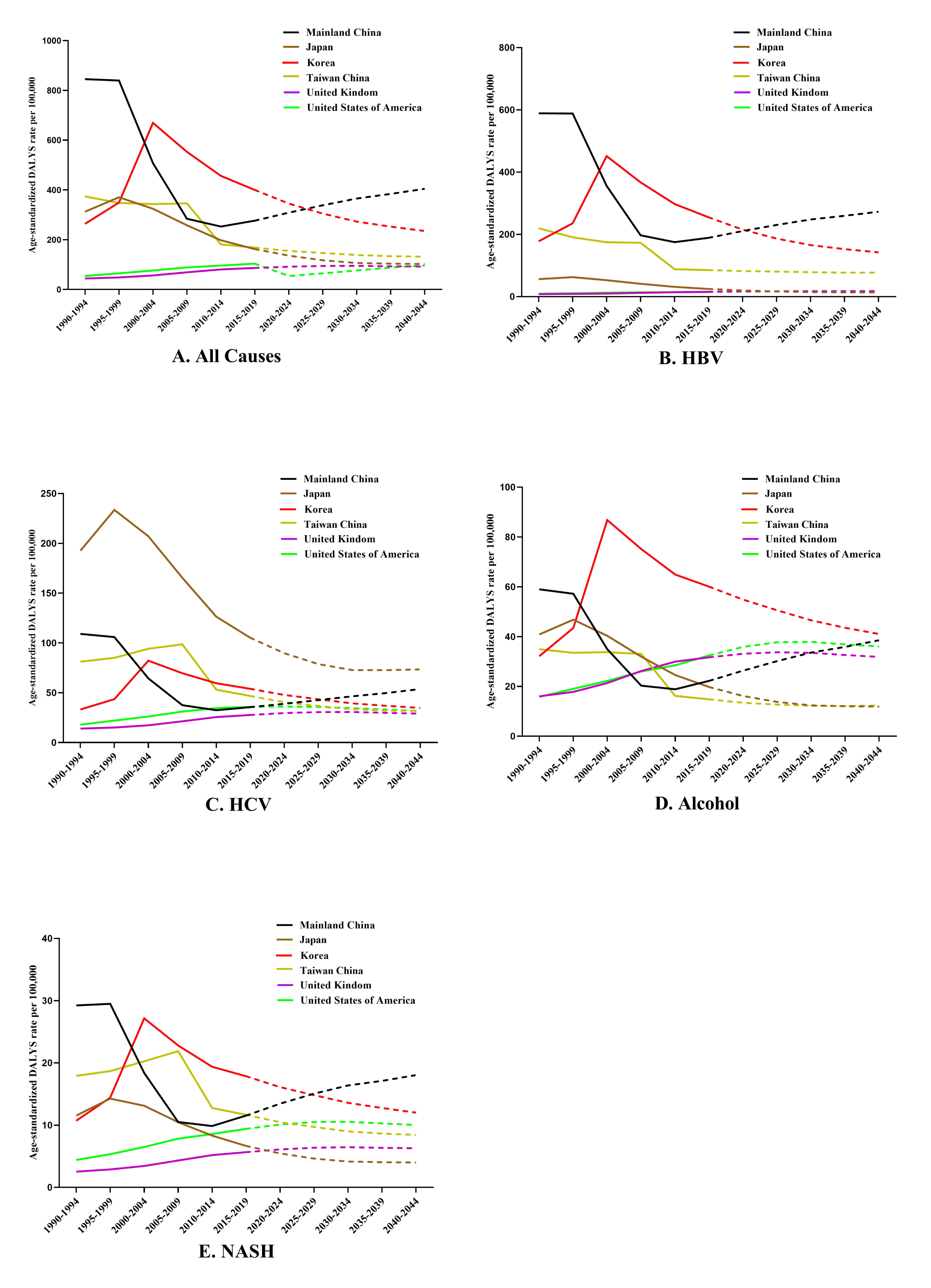
Trends in liver cancer DALYS through 1990 to 2045: observed (solid lines) and predicted rates (dashed lines). Age-standardized to world population per 100,000. DALYS: Disability Adjusted Life Years.

## Discussion

In this study, we demonstrated the highest incidence of liver cancer was observed in Mainland China in 1990 at 28.1 per 100,000 and then declined considerably and ended at 10.6 per 100,000 in 2015. Unexpectedly, for the first time, we found that the liver cancer burden has started to increase again in Mainland China since 2015, unlike the continuous decline in other East Asian areas. Moreover, we predicted that it will continue to increase for the next 25 years and remain 14.8 per 100,000 in 2044, due to the remaining hepatitis-related cancer burden and increase in alcohol-related and NASH-related liver cancers. This alarming projection may imply immediate public health action in China.

Consistent with previous studies(4, 5, 22), the incidence and mortality rates of liver cancer in the current study decreased in East Asia in the past few decades. The decline in liver cancer burden among East Asia areas were clearly attributed to the efforts in preventing hepatitis virus infection. Japanese government launched national projects for HCV blood test since 1989 and provided interferon therapy for HCV-positive adults for five years since 2002(23). Consequently, the incidence of liver cancer in Japan peaked in 2000-2004 and decreased thereafter(24). We also found the plateaued incidence of liver cancer between 2000-2004 in this study and decreased by 78% between 2004 and 2044 in Japan (**Figure 1A**). Similarly, the government of Taiwan China and South Korea launched universal HBV vaccination programs in 1984 and 1995, respectively(24). Subsequently, HBV-related liver cancer incidence in young adults was reported to decrease significantly(25). Followed these East Asia areas, the government of Mainland China has issued a “blood donation law” since 1998, which regulated blood donation and prevented the blood transmission of HBV. Furthermore, a nationwide policy of free HBV vaccination for all newborns and children has been universally implemented since 2002(26, 27). These efforts markedly reduced the HBV surface antigen (HBsAg) carrier rates in Chinese populations from 10.22% to 3.59% (unvaccinated vs. vaccinated population)(28, 29). Along with these effective public health policies, our results showed that the hepatitis-related liver cancer burden in Mainland China sharply decreased from 23.1 per 100,000 (18.6 per 100,000 for HBV; 6.9 per 100,000 for HCV) in 1990 to 8.6 per 100,000 (6.9 per 100,000 for HBV; 1.7 per 100,000 for HCV) in 2015, following what had happened in Japan, Korea, and the rest of East Asia (**Table 1**).

However, unlike the consistently declined liver cancer burden in other East Asia areas, the hepatitis-related cancer burden in Mainland China remained stable between 2015- 2019 and projected to increase till 2044 in our study (9.4 per 100,000 for HBV; 2.3 per 100,000 for HCV). As previously reported in South Korea and Taiwan China, the “lag effect” might account for the elevated disease burden in Mainland China, where the incidence of liver cancer among elderly hepatitis reservoirs remains high(15, 30). Yue *et al.* found that the rate of elderly HCC patients increased by 55.9% in Mainland China over 3 decades, related deaths increased by 39.9%, and the corresponding disease burden increased by 31.4%(31). Moreover, less than one-third of Chinese adults were reported to establish vaccine-mediated immunity for hepatitis virus, and these high-risk people might eventually contribute to the elevated disease burden of liver cancer in China(32). Taken together, it was necessary for the Chinese government to implement a nationwide catch-up vaccination policy in the adult population. In addition, active treatment policies for hepatitis reservoirs should also be enhanced, while approximately 28 million HBV carriers and 200,000 new HCV cases need treatment in Mainland China(33, 34).

In addition to the remaining hepatitis-related cancer burden, emerging challenges such as alcohol and NASH might be responsible for the rapid increasing liver cancer burden in Mainland China. In the current study, increased liver cancer burden in the UK and US was widely observed, where alcohol and NASH rather than hepatitis virus played major roles in contrast to East Asia(12, 35). Interestingly, the changing patterns of alcohol- and NASH-related liver cancer incidence were found similar among Mainland China (0.93 to 1.51 per 100,000 for alcohol; 0.5 to 0.73 per 100,000 for NASH) and Western countries in our projection (**Figure 1D, E**), which indicated the changing etiologies of liver cancer in Mainland China(36). Over decades, the alcohol consumption almost doubled from 4.1 liters to 7.2 liters per person in Mainland China, and 22.7% of Chinese adults had engaged in heavy episodic drinking(37). Similarly, overweight and obesity have increased rapidly in the past four decades in China, with rates estimated at 11.1% for overweight and 7.9% for obesity in children and adolescents(17), total liver cancer cases were estimated to increase 48% from 2016 to 2030, considering the emerging NAFLD- and NASH-related cases(38). Consistent with the prevalence of alcohol abuse and overweight, we predicted that the incidence of alcohol- and NASH-related liver cancer increase by 62% and 46%, respectively, in Mainland China during 2020-2044.

Therefore, it is vital to prevent alcohol abuse and obesity from individual and government aspects. For individuals, interventions should be provided by trained staff to initiate changes in risky binge drinking once individuals are screened or assessed for problematic alcohol use(39). In addition, increasing alcohol taxes, the alcohol policy environment, including liquor licensing laws, accessibility restrictions, and minimum legal drinking age laws, should be enhanced by the government to reduce alcohol use at the population level(40). Healthy diet, increased physical activity, and weight loss should be encouraged for overweight individuals(41). Of importance, universal education for obesity control must be provided to better inform the public in Mainland China(42), in particular, promoting changes in lifestyle and diet for Chinese children to help relieve the future obesity burden(43).

Our study is population-based and incorporates high-quality data from the GBD study, which reliably collects and classifies cancer cases at the global level. Nevertheless, several limitations should be noted. First, some important etiologies, such as aflatoxin B1, tobacco smoking, and multi-etiological liver cancer, were not discussed due to the scarcity of data in the GBD database. Second, the NOREPRED model performs projection based on the assumption that trends observed in the past will continue in the future, which may not be the case. In addition, data from the GBD database may not be detailed enough to make precise predictions. For example, we recently found an overall decline in the disease burden of hepatocellular carcinoma in the US population using the SEER database, in contrast to the currently increasing trends(44); besides, An *et al.* observed the overall decline in HCC incidence among Chinese population from national database(16), thus, our findings must be interpreted with caution. Third, GBD studies provide metrics rather than original data; however, the GBD database is an unparalleled resource for the overall prediction and comparison of disease trends among countries and regions. Although the prediction should be reexamined comprehensively using more detailed and original data from the national registry of China, the possible resurgence of liver cancer burden found in our study is alarming. Previous studies have predicted that the disease burden will continue to decline in Mainland China(45). Our timely prediction is important in urging the Chinese society to rethink and take action to prevent the emerging resurgence of the liver cancer burden.

## Conclusion

Unexpectedly, the current study predicted that the declined liver cancer burden in Mainland China will reverse and increase again through 2044, due to remaining hepatitis-related cancer burden and changing prevalence of alcohol and NASH in Mainland China. The increasing trend we predict requires vigilance from the public health care providers and policymakers in China and calls for enhanced prevention and treatment of hepatitis reservoirs. Besides, we specifically propose the encouragement of enhanced lifestyle modifications, such as drinking cessation and obesity prevention, especially for the younger generation in Mainland China. Our analyses are informative and may guide further action for control the liver cancer burden in Mainland China.

## Abbreviations

ASDR: age-standardized disability-adjusted life-years rate; ALD: alcoholic liver disease; ASIR: age-standardized incidence rate; ASMR: age-standardized mortality rate; DAA: Direct-acting antiviral; DALYS: disability-adjusted life-years; GBD: the Global Burden of Disease study; HBV: Hepatitis B virus; HBsAg: Hepatitis B virus surface antigen; HCV: Hepatitis C virus; NAFLD: nonalcoholic fatty liver disease; NASH: nonalcoholic steatohepatitis; UK: the United Kingdom; US: the United States of America.

## Data Availability

The datasets generated for this study are available on request to the corresponding author.

## Acknowledgement

We sincerely thank the work performed by the Global Burden of Disease Study 2019 collaborators for providing this invaluable database.

## Ethical approval statement

Ethical approval and consent were not required as this study was based on publicly available data.

## Patient consent

Patient consents were not required in accordance with local institution guidelines because the study is a retrospective database research in nature, there was no direct patient contact. Institutional Review Board approval was not required according to our institution policy.

## Author contribution

ZF and LZ: guarantor of the article. ZF, LZ, JH, TT and CC: conception/design. JH, CC, TT, and WL: collection and/or assembly of data. JH, CC, TT, and RC: data analysis and interpretation. JH, CC, RC, SL, HD and TJ: manuscript writing. JH, CC, TT, WL, RC, SL, HD, TJ, and ZF: final approval of manuscript. The work reported in the paper has been performed by the authors, unless clearly specified in the text.

## Data sharing statement

The datasets generated for this study are available on request to the corresponding author.

